# Factors associated with shisha smoking: a cross-sectional telephone-based survey among general population adults in Nigeria

**DOI:** 10.1101/2024.02.27.24303237

**Authors:** Noreen Dadirai Mdege, Sharon Ogolla, Seember Joy Ali, Aminata Camara, Malau Mangai Toma, Emmanuel Agbons Abraham, Victor Olufolahan Lasebikan

## Abstract

**Background:** This is the first national-level study to identify factors associated with shisha smoking among general population adults in Nigeria where high prevalence rates have been reported.

**Methods:** We conducted a telephone-based, cross-sectional survey between 28th July and 11th September 2022 in 12 states. We performed logistic regression analysis, with adjusted odds ratios (aORs) and 95% confidence intervals (CI) as the measures of association.

**Findings:** We surveyed 1278 individuals: 611 who currently smoked shisha and 667 who did not. The following increased the likelihood of being a person who currently smokes shisha: being a person who currently smokes cigarettes (aOR=5.54, 95% CI 2.57 to 11.90) or consumes alcohol (aOR=3.46, 95% CI 1.91 to 6.28); and having a family member (aOR=2.32, 95% CI 1.23 to 4.40), or one (aOR=22.81, 95% CI 9.99 to 52.06) or more (aOR=78.85, 95% CI 22.50 to 276.33) close friends who smoke shisha. The following reduced the likelihood of being a person who currently smokes shisha: being older (aOR=0.92, 95% CI 0.89 to 0.95) and screening positive for possible generalised anxiety disorder (aOR=0.60 95% CI 0.41 to 0.88). The following did not have an effect on current shisha smoking status: sex, level of education, employment status, household wealth, religion, rural/urban residence, perceived stress score, and screening positive/negative for possible major depressive disorder.

**Conclusions:** Strategies to curb shisha smoking need to account for the associated social and behavioural factors, including age, cigarette smoking, alcohol consumption, and having family members or close friends who smoke shisha.

**KEY MESSAGES:** *What is already known on this topic:*

- Shisha smoking is a growing public health threat worldwide, including in Africa, but there is a lack of general population, national-level studies that identify the factors associated with shisha smoking in this context.

*What this study adds:*

- The study suggests that the following increase the likelihood of being a person who smokes shisha: being a person who smokes cigarettes or drinks alcohol, having family members or close friends who smoke shisha, and being a younger adult.
- Females seem to be as likely to smoke shisha as their male counterparts in this context.
- However, there are potential sex differences in the factors associated with shisha smoking, for example, the effects of household wealth, employment status, having family members who smoke shisha, and mental health
- The relationship between shisha smoking and mental health might be different from that which is well-known between smoking other tobacco products such as cigarettes and mental health.

*How this study might affect research, practice or policy:*

- Interventions to curb shisha smoking need to account for the factors associated with the behaviour, sex differences, as well as differences that might exist between shisha smoking behaviour and the smoking of other tobacco products.

## INTRODUCTION

Shisha smoking is becoming more common worldwide (1–6), despite its negative health consequences such as cancers, lung and cardiovascular problems, infertility, and adverse pregnancy outcomes such as low birth weight in babies born to mothers who smoke shisha (7, 8). The smoke contains carcinogens and other toxicants such as nitric oxide and heavy metals, and high levels of carbon monoxide from the burning charcoal (9, 10). For Nigeria, studies among young people have reported high shisha smoking prevalence rates ranging from 3% to 7% (4, 11, 12). Nigeria has made a number of commitments to addressing tobacco use, including shisha smoking, through evidence-informed interventions. These include signing and ratifying the World Health Organization Framework Convention on Tobacco Control (WHO FCTC) (13); and signing into law the National Tobacco Control (NTC) Act in 2015 and the NTC implementing Regulations in 2019 (3–5, 14, 15). Achieving the country’s tobacco control objectives is, however, highly dependent on the availability of context-specific evidence to inform policy decisions.

Unfortunately, population-based, national-level data on shisha smoking is scarce in Nigeria, including on the prevalence, smoking patterns and factors that influence the behaviour. The few studies that exist focus on specific populations such as secondary school and university students, medical professionals or nightclub patrons, and cover a few geographical areas (4, 5, 11, 12, 16–20). For example, we identified only five studies in Nigeria on factors associated with shisha smoking (4, 5, 16, 20, 21), and none of them were conducted in the general population: four were among secondary school and university students (4, 16, 20, 21), whilst one was among nightclub patrons (5). Three of these five studies were conducted solely in Oyo state (5, 16, 21), with one study each for Rivers state (20) and Lagos state (4). They, therefore, only cover three out of Nigeria’s 36 states and Federal Capital Territory (FCT). These shortcomings severely limit the generalizability of the evidence and its usefulness for policy decisions. We addressed this existing evidence gap by investigating the factors associated with shisha smoking in a large sample of general population adults in 12 states across all six geopolitical zones in Nigeria. This evidence can inform decisions of population- or individual-level interventions to reduce shisha smoking in Nigeria and other similar contexts.

## METHODS

### Study design and sites

This was a telephone-based, cross-sectional, quantitative survey conducted between 28th July and 11th September 2022. We conducted the study in states where the prevalence of shisha smoking was high in order to increase the chances of enrolling enough people who smoke shisha. In the absence of national- or state-level data on shisha smoking in Nigeria, we considered urbanisation rate, characterised by the presence of bars, modern restaurants, and social clubs, as well as a more educated and younger population as a proxy for shisha smoking prevalence (5, 22). Using data from Nigeria’s Demographic and Health Survey conducted in 2018 (23), we selected the two most urbanised states/FCT in each of the six geopolitical regions for a total of 12 states/FCT. If, for a geopolitical zone, the most urbanised state was judged as not feasible for study implementation, for example, due to recurring insecurity challenges, it was replaced by the state with the next highest urbanisation rate. Each participating state had its enumeration areas (EAs) stratified into rural or urban EAs. Seven urban EAs and three rural EAs were then randomly selected in each participating state for data collection.

### Study sample

The eligibility criteria for study participation were as follows: 1) aged ≥18 years; 2) a resident in one of the study areas; 3) having a phone number listed in the 2018/19 Nigeria Living Standard Surveys (NLSS) sample frame; and 4) no other household member already participating in the study. The 2018/19 NLSS was an in-person survey where phone numbers of up to three household members were collected, and the sample frame is representative at national-, zonal-, and state-level including the FCT. Other available information includes the state, local government area, rural/urban status, and EA within which each household in the frame belongs, and other background information of respondents.

We aimed to call 8000 phone numbers in our sampling frame. Assuming a 15% response rate, we would be able to achieve our target sample size of 1200 needed to make the comparison between our two groups of interest (i.e., 600 people who currently smoked shisha and 600 who did not smoke shisha currently), within a 3% margin of error with associated 95% confidence levels (24). Using simulations and the number of events per variable approach, this sample size was adequately powered to produce accurate estimates that represent the target population parameters (24, 25). We aimed to enrol 10 individuals (five who currently smoked shisha and five who did not smoke shisha currently; about three females and seven males) from each of the 120 participating EAs.

### Participant recruitment and enrolment

For each participating EA, phone numbers were randomly drawn from the NLSS sample frame and called until 10 eligible individuals had been enrolled and completed the questionnaire. If the call was answered, the respondent was given brief information on the study aims and objectives and screened for eligibility if they were willing to continue with the conversation. If they were eligible, they were given more detailed information about the study including what participation would involve, the types of information collection, how the information would be used, time taken, confidentiality, voluntary nature of participation and anonymity of their responses. Verbal consent was obtained before data collection commenced. Any participants who consented to the survey but were unable to participate at that time were asked for an appointment at a time convenient to them. For some EAs, random selection of phone numbers could not yield enough people who currently smoked shisha, and this was complemented by recruitment from adjacent EAs in the same state or snowball sampling.

### Measures

The development of the questionnaire, and the questions included, has been described in detail elsewhere (26). Briefly, the questionnaire was informed by literature reviews, the Theory of Planned Behaviour (27–31), and a qualitative study that preceded this survey (26). The dependent variable, current shisha smoking status, was self-reported from two questions: 1) *“Have you ever smoked shisha, even one or two puffs?”*, plus 2) *“Do you currently smoke shisha on a daily basis, less than daily or not at all?”*. Those who responded “no” to the first, and those responding “not at all” to the second question were classified as people who did not smoke shisha currently. Those who responded “yes” to the first, and “daily” or “less than daily” to the second question were classified as people who currently smoked shisha. Independent variables included demographic and socioeconomic characteristics age, sex, rural/urban residence, level of education, religion, employment status, and asset-based household wealth based on household ownership of 12 assets (23, 32). The following were also included as independent variables as they have been found to be associated with tobacco smoking: subjective norm-related variables, specifically having family members or close friends who smoke shisha; current cigarette smoking status (smoked cigarettes/not in the last year); current alcohol consumption status (drunk alcohol/not in the last year); and stress, anxiety and depression (32–37). Perceived stress was measured using the 4-item Perceived Stress Scale (PSS-4) (38–40); possible major depressive disorder using the Patient Health Questionnaire-2 (PHQ-2) (41); and possible generalized anxiety disorder using the General Anxiety Disorders-2 (GAD-2) (42).

### Data collection

Data was collected by 23 enumerators who had received a 5-day training, using Computer Assisted Telephone Interviews (CATI). The piloted questionnaire was programmed using the SurveyCTO software which offers end-to-end encryption of data (43), on password-protected android tablets. The questionnaire was administered in English as well as the four major local languages in Nigeria, i.e., Hausa, Igbo, Yoruba and Pidgin English. The training of enumerators covered the questionnaire content and administration, safeguarding principles, research ethics and other relevant topics. The training involved classroom training, role-playing and field practice and testing using the CATI devices. Data was collected from a call centre and the questionnaire took up to 35 minutes to administer. Each participant received five hundred Naira (∼1.2 USD at the time of the study) worth of airtime as compensation for their time.

### Data analysis

We summarised and presented categorical variables as percentages and continuous variables as means and standard deviations (SD). We used adjusted odds ratios (ORs), with 95% confidence intervals (CI), as the measures of association. Bivariate analysis was conducted between each independent variable and the dependent variable, and all independent variables with a p-value < 0.2, and those that were known, theoretically or empirically, to be associated with smoking status were included in multivariate analysis (36). The following variables were, therefore, included in the final multivariate model: age, sex, rural/urban residence, level of education, religion, household wealth, employment status, having family members who smoke shisha, the number of closest friends who smoke shisha, current cigarette smoking status, current alcohol consumption status, stress, depression and anxiety. Sample weights were applied, with the weight of each observation being equal to the inverse of the probability of selection. For competing multivariate analysis models, the best model was determined using Akaike’s information criteria (AIC), while the Hosmer–Lemeshow test was run to test the model’s goodness-of-fit. Variance Inflation Factor (VIF) analysis was used to test for multicollinearity in the regression models. We conducted subgroup analysis by sex. Analyses were conducted in STATA version 18 (44).

## RESULTS

### Participant characteristics

7733 phone numbers were called from which 1278 (16.5%) survey questionnaires were successfully completed (Figure 1). 611 (47.8%) survey participants were people who currently smoked shisha and 667 (52.2%) were people who did not smoke shisha currently. 12.4% of the people who currently smoked shisha smoked it daily and 87.6% smoked it less than daily. Among the people who did not smoke shisha currently, 88.8% (n=592) had never smoked shisha and 11.2% (n=75) had smoked it in the past.

**Figure 1:**
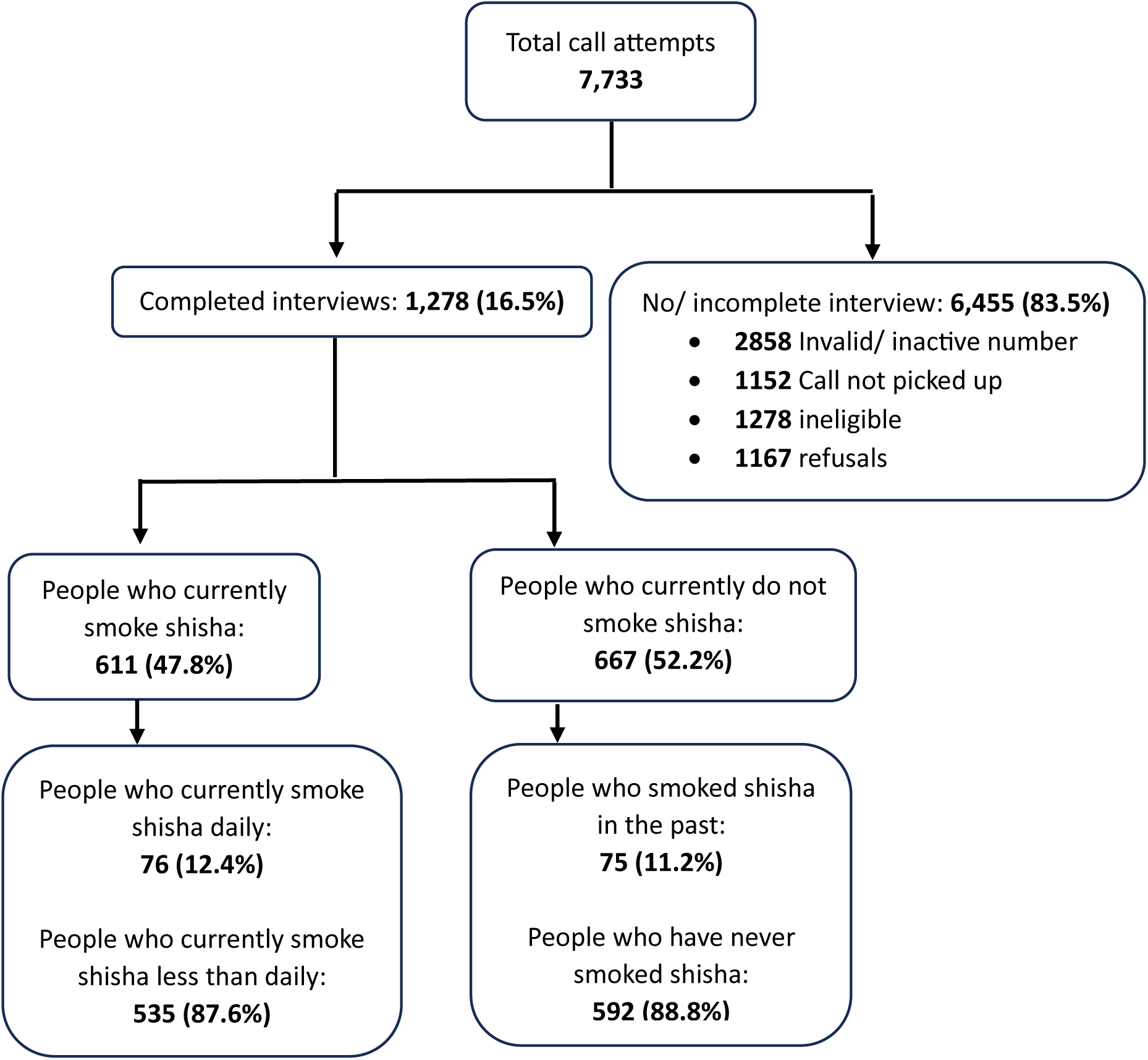
Flow of participants in the study.

Figure 2 shows the distribution of current shisha smoking status by geopolitical zone. It also shows the states where data was collected from in each geopolitical zone.

**Figure 2:**
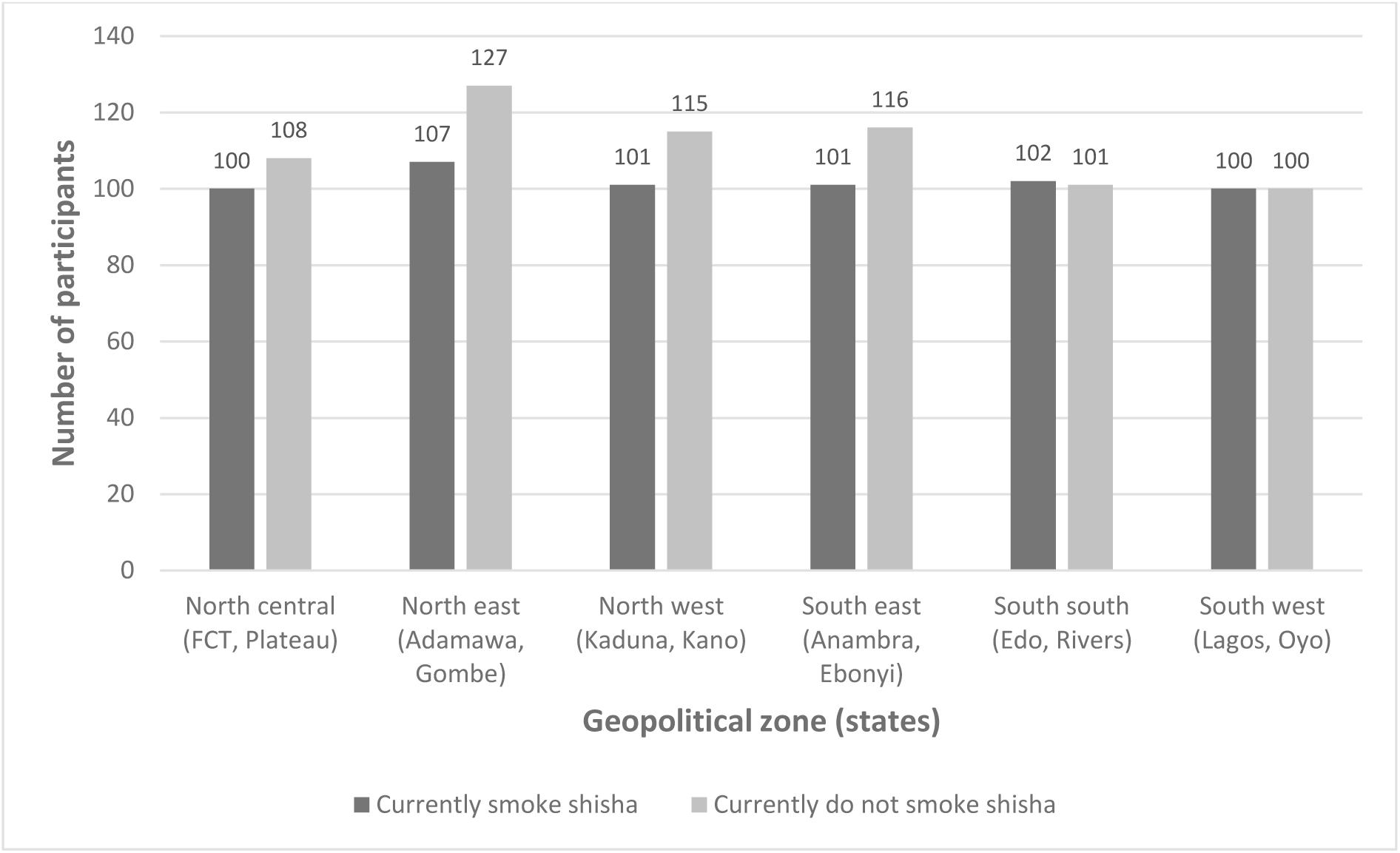
Number of participants by current shisha smoking status and geopolitical zone.

The average age of the survey participants was 32.3 years, and 69% were male with 31% being female (Table 1). 72% and 28% were from urban and rural areas, respectively. Over 90% of participants had a minimum of secondary education. The majority of participants (∼73%) were employed. Distribution by religion and wealth are also in Table 1. From Table 2, approximately 16% of participants had at least one family member who smokes shisha, and approximately 60% had at least one closest friend who smokes shisha. 12% of the participants currently smoked cigarettes whilst 47% currently consumed alcohol. The mean PSS-4 score was 7.0 (SD=2.5). 41% of the study participants screened positive for possible major depressive disorder, and 39% screened positive for possible generalized anxiety disorder. Characteristics by current shisha smoking status are also provided in Tables 1 and 2.

**Table 1:** General demographic characteristics and their association with current shisha smoking status.

| **Variable** | **Overall**<br>**(n=1,278)** | **Currently smoke**<br>**shisha (n=611)** | **Currently do not smoke**<br>**shisha (n=667)** | **cOR (95% Confidence**<br>**Interval (CI))** | **aOR (95% CI)** |
| --- | --- | --- | --- | --- | --- |
| **Age mean (sd)** | 32.3 (9.7) | 28.8 (6.0) | 35.6 (11.1) | 0.91 (0.89 to 0.94) | **0.92 (0.89 to 0.95)** |
| **Sex: n (%)** |  |  |  |  |  |
| Female | 394 (30.8) | 145 (23.7) | 249 (37.3) | 1 | 1 |
| Male | 884 (69.2) | 466 (76.3) | 418 (62.7) | 1.92 (1.39 to 2.65) | 0.92 (0.56 to 1.52) |
| **Urban/rural: n (%)** |  |  |  |  |  |
| Rural | 361 (28.2) | 140 (22.9) | 221 (33.1) | 1 | 1 |
| Urban | 917 (71.8) | 471 (77.1) | 446 (66.9) | 1.66 (0.99 to 2.80) | 1.26 (0.62 to 2.54) |
| **Level of education: n (%)** |  |  |  |  |  |
| No education | 44 (3.5) | 4 (0.7) | 40 (6.0) | 1 | 1 |
| Primary | 73 (5.8) | 8 (1.3) | 65 (9.7) | 1.29 (0.36 to 4.64) | 0.47 (0.15 to 1.50) |
| ≥ Secondary | 1161 (91.7) | 599 (90.0) | 562 (84.3) | 10.47(3.67 to 29.87) | 0.54 (0.17 to 1.65) |
| **Religion: n (%)** |  |  |  |  |  |
| Christianity | 803 (62.8) | 417 (68.2) | 386 (57.9) | 1 | 1 |
| Islam | 450 (35.2) | 182 (29.8) | 268 (40.2) | 0.62 (0.45 to 0.85) | 0.99 (0.67 to 1.47) |
| None | 25 (2.0) | 12 (2.0) | 13 (1.9) | 0.94 (0.29 to 3.03) | 0.75 (0.17 to 3.33) |
| **Wealth quintiles: n (%)** |  |  |  |  |  |
| Poorest | 257 (20.1) | 184 (30.1) | 73 (11.0) | 1 | 1 |
| Poorer | 350 (27.4) | 200 (32.7) | 150 (22.5) | 0.53 (0.41 to 0.69) | 0.82 (0.49 to 1.36) |
| Middle | 165 (13.9) | 84 (13.8) | 81 (12.1) | 0.44 (0.29 to 0.68) | 0.71 (0.34 to 1.49) |
| Richer | 266 (20.8) | 89 (14.6) | 177 (26.5) | 0.21 (0.12 to 0.35) | **0.41 (0.18 to 0.97)** |
| Richest | 240 (18.8) | 54 (8.8) | 186 (27.9) | 0.12 (0.08 to 0.19) | 0.49 (0.15 to 1.57) |
| **Employment status: n (%)** |  |  |  |  |  |
| Unemployed | 108 (8.6) | 32 (5.2) | 76 (11.4) | 1 | 1 |
| Employed | 928 (72.6) | 440 (72.0) | 488 (73.2) | 2.21 (0.98 to 5.01) | 1.72 (0.78 to 3.81) |
| Student | 161 (12.6) | 106 (17.4) | 55 (8.2) | 4.62 (2.16 to 9.90) | **2.21 (1.38 to 3.53)** |
| Apprentice | 81 (6.3) | 33 (5.4) | 48 (7.2) | 1.67 (1.04 to 2.69) | 1.45 (0.45 to 4.68) |

**Table 2:**
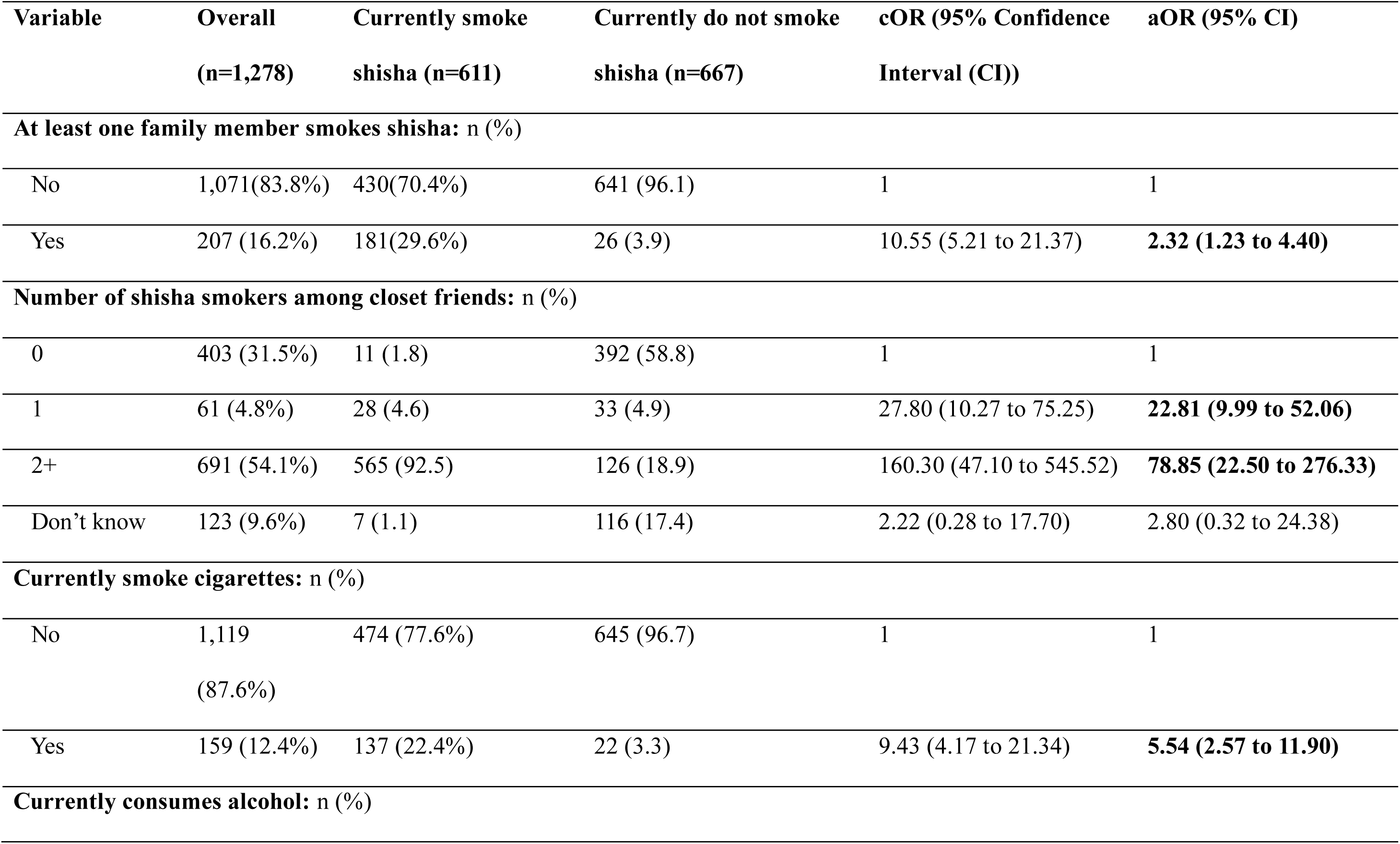
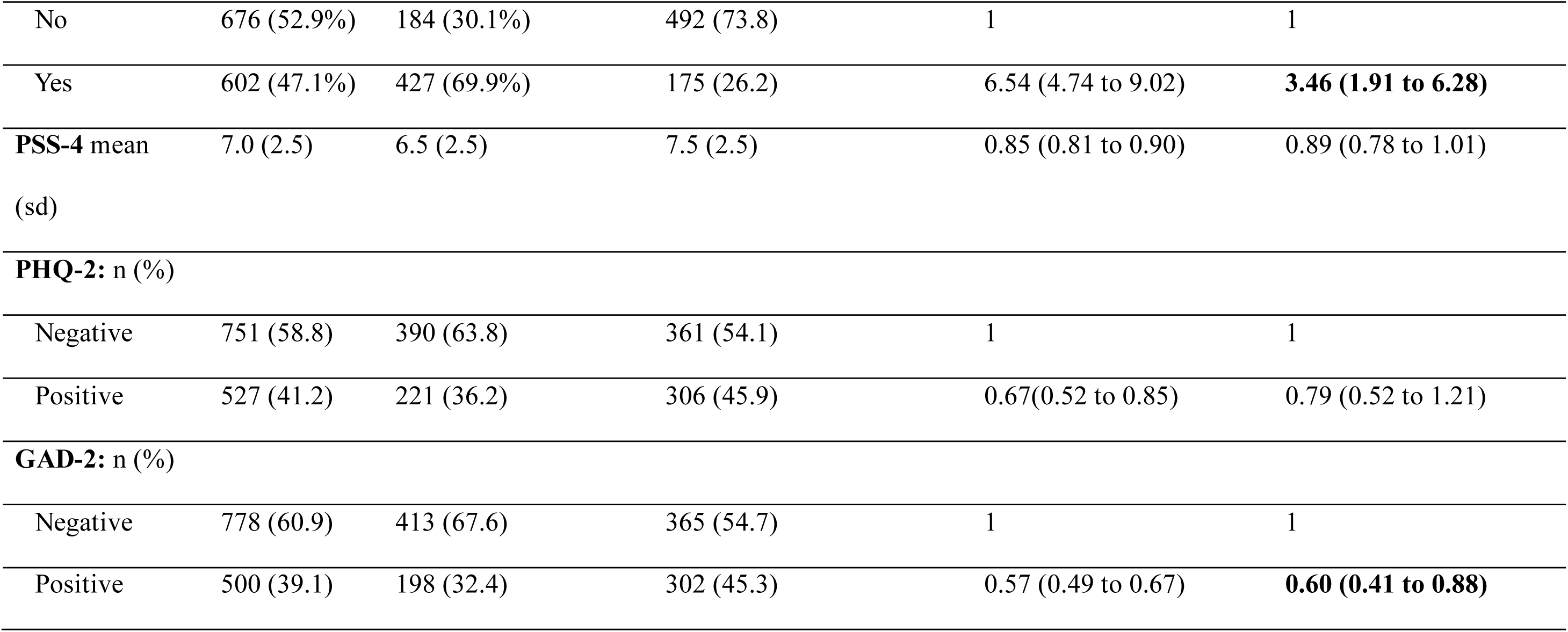
Cigarette and alcohol use, mental health, shisha smoking by family or friends, and association with current shisha smoking status.

### Factors associated with current shisha smoking status

The likelihood of being a person who currently smokes shisha decreased with increasing age (aOR=0.92, 95% CI 0.89 to 0.95, p=0.000) (Table 1). Whilst there was no clear relationship between household wealth and current shisha smoking status across all wealth quintiles, those who belonged to the “richer” quintile were less likely to be people who currently smoke shisha compared to those who belonged to the “poorest” quintile (aOR=0.41, 95% CI 0.18 to 0.97, p=0.043). Those who were students were more likely to be people who currently smoke shisha than those who were currently unemployed (aOR=2.21, 95% CI 1.38 to 3.53, p=0.004). There was no statistically significant association between sex, level of education, religion, or rural/urban residence and current shisha smoking status.

Those who currently smoked cigarettes had higher odds of being people who currently smoke shisha than those who did not currently smoke cigarettes (aOR=5.54, 95% CI 2.57 to 11.89, p=0.001) (Table 2). Those who currently consumed alcohol were more likely to be people who currently smoke shisha than those who did not currently consume alcohol (aOR=3.46, 95% CI 1.91 to 6.28, p=0.001). Those who had a family member who smokes shisha had higher likelihood of being people who currently smoke shisha than those who did not have any family member who smoke shisha (aOR=2.32, 95% CI 1.23 to 4.40, p=0.015). Similarly, those who had one (aOR=22.81, 95% CI 9.99 to 52.06, p=0.000) or more (aOR=78.85, 95% CI 22.50 to 276.33, p=0000) closest friends who smoke shisha had higher odds of being people who currently smoke shisha than those without any closest friends who smoke shisha. Those who screened positive for possible generalised anxiety disorder had lower odds of being people who currently smoke shisha than those who screened negative (aOR=0.60, 95% CI 0.41 to 0.88, p=0.014). Overall, the study findings did not show statistically significant associations between perceived stress scores or screening positive/negative for possible major depressive disorder and current shisha smoking status.

The results for males were similar to the overall results with respect to the association between the following and current shisha smoking status: age, current cigarette smoking status, current alcohol consumer status, having a family member who smokes shisha, having one or more closest friends who smoke shisha, screening positive/negative for possible major depressive disorder, rural/urban residence, level of education, religion and household wealth (Supplemental material 1). However, for employment status, both students (aOR=3.85, 95% CI 1.43 to 10.41, p=0.013) and those who were employed (aOR=2.53, 95% CI 1.09 to 5.89, p=0.034) were more likely to be people who currently smoke shisha than those who were unemployed. In addition, the likelihood of being a person who currently smokes shisha decreased with increasing perceived stress scores (PSS-4) (aOR=0.85, 95% CI 0.72 to 0.99, p=0.044); and there was no statistically significant association between screening positive/negative for possible generalised anxiety disorder and current shisha smoking status.

The results for females were similar to the overall results with respect to the association between the following and current shisha smoking status: age, current alcohol consumer status, having one or more closest friends who smoke shisha, perceived stress score, rural/urban residence, level of education and religion (Supplemental material 1). However, the model fitted for females perfectly predicted the relationship between current cigarette smoking status and shisha smoking status (i.e. odds ratio=1): i.e., all females who currently smoked cigarettes also currently smoked shisha and all females who did not currently smoke cigarettes also did not currently smoke shisha. Contrary to the overall results and those observed for males, there was no statistically significant association between household wealth, employment status or having a family member who smokes shisha and current shisha smoking status. There was also no statistically significant association between screening positive for possible generalised anxiety disorder and current shisha smoking status. However, those who screened positive for possible major depressive disorder were less likely to be people who currently smoke shisha than those who screened negative (aOR=0.36, 95% CI 0.17 to 0.81, p=0.019).

## DISCUSSION

We found that being a person who currently smokes cigarettes, being a person who currently consumes alcohol, having a family member who smokes shisha, or having closest friends who smoke shisha increased the likelihood of being a person who currently smokes shisha than not being/ not having. An increase in age reduced the likelihood of being a person who currently smokes shisha, and those who screened positive for possible generalised anxiety disorder were less likely to be people who currently smoke shisha than those who screened negative. There was no statistically significant association between the following and current shisha smoking status: sex, level of education, religion, rural/urban residence, perceived stress score, and screening positive/negative for possible major depressive disorder. Although there was no clear relationship between household wealth and current shisha smoking status, those who were in the ‘richer’ category were more likely to be people who currently smoke shisha than those in the poorest category. Students were more likely to be people who currently smoke shisha than the unemployed.

Overall, our results align with those from other studies that have found a statistically significant, positive association between cigarette smoking status, alcohol consumption status, or shisha smoking among family members or close friends and being a person who currently smokes shisha (4, 5, 45, 46). Other studies have also found a negative association between age and shisha smoking, with younger individuals being more likely to be people who currently smoked shisha than older individuals (1, 47). However, the three studies in Nigeria that have looked at age did not find any statistically significant relationship with shisha smoking status. Two of these studies were in much younger populations of secondary school and/or university students (mean ages of 20-23 years versus 32 years for our general population sample) (4, 20). The remaining one was also in a different population, i.e., nightclub patrons (5). Many studies report that males are more likely to be people who currently smoke shisha than females (47, 48). However, as with our study, most studies in Nigeria have found no statistically significant difference between males and females in the likelihood of being a person who currently smokes shisha (4, 5, 20). We identified only one study in Nigeria suggesting that males were more likely to be people who currently smoke shisha than females (16). From our previous report using qualitative interviews with 78 people who currently smoked shisha and 611 people who currently smoked shisha from the study being reported here, although shisha smoking was perceived as generally socially unacceptable in Nigeria, it was perceived as more accepted by society, and more comfortable to females when compared to cigarette smoking (26). This makes it plausible that, unlike other smoked tobacco products such as cigarettes that are more commonly used by males than females, shisha smoking could be as common in females as it is in males in Nigeria. However, there is currently no general population, national-level data to confirm or refute whether this is indeed the case.

Although there is overwhelming evidence of a significant positive association between tobacco smoking and mental health problems such as anxiety, tension and depression (49, 50), we did not observe this relationship in our study. Instead, we found that those who screened positive for possible generalised anxiety disorder were less likely to be people who currently smoke shisha than those who screened negative. We have previously reported that our subsample of 611 people who currently smoked shisha, and the 78 people who currently smoked shisha we had qualitative interviews with, associated shisha smoking with pleasure and fun, and mostly smoked shisha with close friends in places that provided food and entertainment (26). Only 13% of the 611 people who smoked shisha reported smoking shisha to cope with challenging life situations such as stress (26). This suggests that the relationship between shisha smoking and mental health might be different from that which exists between smoking other smoked tobacco products such as cigarettes and mental health.

There were a number of sex differences in the results. Whilst for males the relationship between household wealth and current shisha smoking status was the same as the overall result, there was no statistically significant relationship for females. Among males, students and those who were employed were more likely to be people who currently smoke shisha than those who were unemployed; but for females, there was no statistically significant relationship between employment status and current shisha smoking status. We also found that for females, having close friends who smoke shisha predicted shisha smoking status, and having family members who smoke shisha did not; but for males, both were predictors. This might stem from the fact that societal views are more negative (and therefore potentially more protective) towards female than males who smoke shisha, which potentially limits the impact of family members’ shisha smoking on female behaviour. In our previous analysis we found that many women who smoked shisha mainly smoked it privately or in safe spaces, particularly among close friends, where there was less likelihood of stigmatisation (26). An increase in the perceived stress score decreased the likelihood of being a person who currently smokes shisha among males, but not among females. Whilst there was no association between screening positive/negative for possible major depressive disorder and current shisha smoking status among males, for females, those who screened positive were less likely to be people who currently smoke shisha than those who screened negative.

Interventions aimed at curbing shisha smoking in Nigeria should consider the influence of family and friends on shisha smoking behaviour, and co-consumption with other tobacco products as well as alcohol. In addition, the interventions should also consider the differences that might exist between other smoked tobacco products and shisha, for example, differences in terms of the influence of sex and mental health status. Sex differences in terms of factors that influence shisha smoking should also be accounted for in order to ensure that the needs of both males and females are catered for. In terms of future research, there is a need for nationally representative data on the prevalence of shisha smoking as well as shisha smoking patterns. The data needs to allow for disaggregation by important demographic characteristics such as sex, age, rural/urban residence and socio-economic status and inform targeted responses where necessary. There is also a need to unpack the differences between shisha smoking behaviour and the smoking of other tobacco products, and the implications of those differences on tobacco control measures.

### Strengths and limitations

The sample was limited to those with phone numbers that were listed in the NLSS, which might restrict the applicability of the findings to the general population in Nigeria. However, the impact of this is reduced by the high phone penetration rates in Nigeria of more than 80% (51), and the fact that NLSS sample frame of households is representative at national-, zonal-, and state-levels including FCTs. In addition, the random selection of 10 EAs in each of the 12 participating states covering all six geographical zones of Nigeria also enhanced the generalizability of our study findings. The snowball sampling introduced in some EAs to get enough people who currently smoke shisha might have introduced selection bias. We anticipate that this did not affect the direction of and of the results. However, this could have potentially led to an over estimation of the aOR for having family or friends who smoke shisha, if those who smoked shisha were more likely to refer other shisha smokers than those who did not smoke shisha. We were able to translate our data collection tools into local languages and also administer the questionnaire and interviews in local languages for those who preferred this which enhanced the accessibility of the study, and hence generalizability.

## CONCLUSIONS

In our study, the following increased the likelihood of shisha smoking: being a person who currently smokes cigarettes or a person who currently consumes alcohol, having family members or closest friends who smoke shisha. Those who were older and those who screened positive for possible generalised anxiety disorder were less likely to be people who currently smoke shisha. Interventions to curb shisha smoking need to take these factors into consideration. In addition, interventions should account for sex differences, as well as differences that might exist between shisha smoking behaviour and the smoking of other tobacco products.

## SUPPLEMENTAL MATERIAL

Supplemental material 1: Factors associated with shisha smoking by sex

Supplemental material 2: STROBE Checklist

## DATA AVAILABILITY STATEMENT

The data used for this manuscript is available in an open access repository at: Development Gateway: An IREX Adventure. (2023). Development Gateway shisha study quantitative data: a cross-sectional survey investigating factors associated with shisha use in Nigeria [Data set]. Zenodo. https://doi.org/10.5281/zenodo.7775958

## CONTRIBUTORS

NDM, SJA, and AC conceptualized and designed the study and the data collection tools. NDM and SO contributed to data analysis and validation. All authors contributed to the interpretation of the results. NDM prepared the first draft of the manuscript, and all the other authors reviewed and provided feedback and edits during the manuscript preparation. All authors approved the final version of the manuscript. NDM is the acting guarantor.

## COMPETING INTERESTS

None declared.

## FUNDING

This work was supported by the Bill & Melinda Gates Foundation (INV-009670).

## DISCLAIMER

The funders had no role in study design, data collection and analysis, decision to publish, or preparation of the manuscript. The findings and conclusions contained within are those of the authors and do not necessarily reflect positions or policies of the Bill & Melinda Gates Foundation.

## ETHICS STATEMENTS

### Patient consent for publication

Not required.

### Ethics approval

Ethics approval for the study was obtained on 21 July 2022 from the National Health Research Ethics Committee, Nigeria (NHREC; approval number NHREC/01/01/2007-21/07/2022). Participants gave informed consent to participate in the study before taking part.

## ACKNOWLEDGMENTS

This study was conducted as part of the Tobacco Control Data Initiative (TCDI). TCDI is a program that covers six African countries (the Democratic Republic of Congo, Ethiopia, Kenya, Nigeria, South Africa, and Zambia) and aims to understand their tobacco data needs, identify existing data, confirm gaps in available tobacco data, collect new data to fill those gaps, and develop tools to enable policymakers to use essential data more effectively to inform tobacco policy. The programme is led by Development Gateway: an IREX Venture (DG), a global non-profit organization that specializes in data for development, in partnership with the University of Cape Town’s Research Unit on the Economics of Excisable Products (REEP). We acknowledge R-DATS Consulting Limited, Abuja, Nigeria who led data collection and analysis on behalf of DG. We express our gratitude to the support provided by key tobacco control stakeholders in Nigeria. Particularly, we would like to thank the Federal Ministry of Health NCD Division and the Tobacco Control Unit, as well as the National Bureau of Statistics for their guidance, support and collaboration during study design and implementation.

## Notes

### Competing Interest Statement

The authors have declared no competing interest.

### Funding Statement

This study was funded by the Bill & Melinda Gates Foundation (INV-009670).

### Author Declarations

The National Health Research Ethics Committee, Nigeria gave ethical approval for this work.

