## Supplemental material 1 for "Factors associated with shisha smoking: a cross-sectional telephone-based survey among general population adults in Nigeria"

**Supplemental material 1: Factors associated with shisha smoking by sex**

| | Males | | Females | |
|---|---|---|---|---|
| **Variable** | **cOR (95% CI)** | **aOR (95% CI)** | **cOR (95% CI)** | **aOR (95% CI)** |
| **Age** | 0.90 (0.88 to 0.93) | **0.92 (0.90 to 0.95)** | 0.89 (0.85 to 0.93) | **0.90 (0.82 to 0.98)** |
| **Urban/rural** | | | | |
| Rural | 1 | 1 | 1 | 1 |
| Urban | 1.64 (0.89 to 3.02) | 1.17 (0.59 to 2.31) | 1.70 (0.86 to 3.38) | 2.30 (0.39 to 13.68) |
| **Level of education** | | | | |
| No education | 1 | 1 | 1 | 1 |
| Primary | 0.83 (0.18 to 3.89) | 0.32 (0.05 to 2.13) | 2.53 (0.24 to 26.78) | 0.93 (0.10 to 8.91) |
| ≥ Secondary | 9.04 (2.64 to 30.87) | 0.46 (0.20 to 1.06) | 10.86 (1.43 to 82.73) | 0.37 (0.02 to 6.71) |
| **Religion** | | | | |
| Christianity | 1 | 1 | 1 | 1 |
| Islam | 0.68 (0.50 to 0.94) | 0.91 (0.65 to 1.27) | 0.32 (0.16 to 0.65) | 0.85 (0.35 to 2.04) |
| None | 0.75 (0.28 to 2.00) | 0.75 (0.18 to 3.22) | 1.70 (0.12 to 24.17) | 0.68 (0.06 to 7.41) |
| **Wealth quintile** | | | | |

| | | | | |
|---|---|---|---|---|
| Poorest | 1 | 1 | 1 | 1 |
| Poorer | 0.49 (0.36 to 0.68) | 0.83 (0.44 to 1.60) | 0.62 (0.39 to 0.98) | 0.78 (0.26 to 2.39) |
| Middle | 0.48 (0.28 to 0.81) | 0.68 (0.34 to 1.37) | 0.20 (0.03 to 1.52) | 0.37 (0.11 to 1.22) |
| Richer | 0.25 (0.16 to 0.39) | **0.39 (0.17 to 0.88)** | 0.12 (0.05 to 0.33) | 0.43 (0.07 to 2.60) |
| Richest | 0.10 (0.06 to 0.17) | 0.35 (0.11 to 1.12) | 0.17 (0.08 to 0.38) | 1.85 (0.12 to 27.69) |
| **Employment status** | | | | |
| Unemployed | 1 | 1 | 1 | 1 |
| Employed | 1.63 (0.54 to 4.89) | **2.53 (1.09 to 5.89)** | 2.02 (0.94 to 4.34) | 0.79 (0.09 to 6.71) |
| Student | 3.73 (1.22 to 11.38) | **3.85 (1.43 to 10.41)** | 5.13 (1.76 to 14.98) | 0.84 (0.18 to 4.06) |
| Apprentice | 1.38 (0.75 to 2.54) | 2.77 (0.77 to 9.96) | 1.47 (0.86 to 2.50) | 0.09 (0.00 to 4.27) |
| **At least one family member smokes shisha** | | | | |
| No | 1 | 1 | 1 | 1 |
| Yes | 9.40 (4.02 to 22.01) | **2.23 (1.16 to 4.29)** | 13.58 (6.31 to 29.25) | **4.91 (0.88 to 27.26)** |
| **Number of shisha smokers among closet friends** | | | | |
| 0 | 1 | 1 | 1 | 1 |

| | | | | |
|---|---|---|---|---|
| 1 | 12.85 (4.15 to 39.79) | **10.81 (4.17 to 28.00)** | 176.40 (46.68 to 666.52) | **289.38 (34.94 to 2396.71)** |
| 2+ | 114.98 (34.74 to 380.55) | **64.17 (21.68 to 189.97)** | 327.47 (42.01 to 2552.79) | **226.58 (20.12 to 2552.09)** |
| Don't know | 2.45 (0.28 to 21.39) | 3.59 (0.27 to 47.66) | 1.05 (0.04 to 27. 35) | 1.98 (0.09 to 44.70) |
| **Currently smoke cigarettes** | | | | |
| No | 1 | 1 | N/A | N/A |
| Yes | 7.20 (3.16 to 16.42) | **4.62 (2.24 to 9.51)** | N/A | N/A |
| **Currently consumes alcohol** | | | | |
| No | 1 | 1 | 1 | 1 |
| Yes | 4.49 (3.27 to 6.16) | **2.82 (1.47 to 5.40)** | 16.73 (7.84 to 35.71) | **8.88 (2.93 to 26.95)** |
| **PSS-4** | 0.83 (0.77 to 0.89) | **0.85 (0.72 to 0.99)** | 0.91 (0.83 to 1.01) | 1.10 (0.93 to 1.30) |
| **PHQ-2** | | | | |
| Negative | 1 | 1 | 1 | 1 |
| Positive | 0.66 (0.47 to 0.92) | 0.85 (0.46 to 1.56) | 0.68 (0.46 to 1.00) | **0.36 (0.17 to 0.81)** |
| **GAD-2** | | | | |

| | | | | |
|---|---|---|---|---|
| Negative | 1 | 1 | 1 | 1 |
| Positive | 0.54 (0.42 to 0.70) | 0.61 (0.32 to 1.14) | 0.66 (0.44 to 0.99) | 0.42 (0.15 to 1.15) |
